## Supplementary Tables for "Emergence of transmissible SARS-CoV-2 variants with decreased sensitivity to antivirals in immunocompromised patients with persistent infections"

**Supplementary Table 1.** Antiviral treatment and viral dynamics in immunocompromised patient cohort.

| Patient ID | Antiviral drug | Treatment day post-diagnosis | Days PCR + | Clade | Days pd | Ct | Accession |
|---|---|---|---|---|---|---|---|
| 1995 | RDV | 1-10 | 28 | 20C | 0 | NA | SAMN40750127 |
| | | | | | 1 | NA | SAMN40750128 |
| | | | | | 6 | NA | SAMN40750128 |
| 2646 | RDV | 15-19 | 32[+] | 21K | 0 | NA | SAMN40750130 |
| | | | | | 26 | 25.6 | SAMN40750131 |
| 10939 | RDV | 2-6, 14-18 | 33[@] | 20C | 13 | 16.5 | SAMN40750132 |
| | | | | | 21 | 25.15 | SAMN40750133 |
| 11595 | RDV | 2-7 | 38 | 20G | 14 | 19.4 | SAMN40750134 |
| | | | | | 20 | 19.25 | SAMN40750135 |
| | | | | | 26 | 20.7 | SAMN40750136 |
| | | | | | 38 | 20.7 | SAMN40750137 |
| 12105 | RDV | 16-20 | 58[@] | 20G | 0 | 14.8 | SAMN40750138 |
| | | | | | 21 | 28.3 | SAMN40750139 |
| 14675 | Paxlovid | 1-5 | 66 | 22C | 0 | 32.3 | SAMN40750140 |
| | RDV | 31-35 | | | 19 | 23.2 | SAMN40750141 |
| | | | | | 24 | 22.5 | SAMN40750142 |
| | | | | | 29 | NA | SAMN40750143 |
| | | | | | 51 | NA | SAMN40750144 |
| 16624 | RDV | 1-5 | 42 | 21K | 0 | NA | SAMN40750145 |
| | | | | | 12 | NA | SAMN40750146 |
| | | | | | 27 | 22.7 | SAMN40750147 |
| 16902 | RDV | 26-37,<br>47-51 | 50[@] | 21K | 0 | NA | SAMN40750148 |
| | | | | | 1 | NA | SAMN40750149 |
| | | | | | 7 | NA | SAMN40750150 |
| | | | | | 11 | NA | SAMN40750151 |
| | | | | | 14 | NA | SAMN40750152 |
| | | | | | 26 | NA | SAMN40750153 |
| | | | | | 28 | 25.1 | SAMN40750154 |
| | | | | | 36 | NA | SAMN40750155 |
| | | | | | 42 | NA | SAMN40750156 |
| | | | | | 47 | 23 | SAMN40750157 |
| 16915 | RDV | 0-2 | 63 | 21K | 0 | NA | SAMN40750158 |
| | | | | | 14 | 20.83 | SAMN40750159 |
| | | | | | 21 | 22.38 | SAMN40750160 |
| | | | | | 35 | 19.8 | SAMN40750161 |
| 17062 | RDV | 0-4 | 72 | 21K | 8 | NA | SAMN40750162 |
| | | | | | 11 | 18.76 | SAMN40750163 |
| 17320 | RDV | 31-33 | 34 | 21K | 30 | 22.4 | SAMN40750170 |
| | | | | | 31 | 21.4 | SAMN40750171 |
| | | | | | 34 | 25.5 | SAMN40750172 |
| 17386 | RDV | 3-7,<br>38-42,<br>108-111,<br>148-152 | 157 | 21K | 104 | 24.8 | SAMN40750173 |
| | | | | | 141 | NA | SAMN40750174 |
| | | | | | 143 | 20.8 | SAMN40750175 |
| 17423 | RDV | 0-2 | 190 | 21K | 0 | NA | SAMN40750176 |
| | | | | | 3 | 21.5 | SAMN40750177 |
| | | | | | 15 | 20.5 | SAMN40750178 |
| | | | | | 32 | 14.5 | SAMN40750179 |
| | | | | | 39 | 15.2 | SAMN40750180 |
| | | | | | 42 | 28.2 | SAMN40750181 |
| | | | | | 53 | 18 | SAMN40750182 |
| | | | | | 60 | 23.4 | SAMN40750183 |
| | | | | | 67 | 25.5 | SAMN40750184 |
| | | | | | 81 | 19.7 | SAMN40750185 |
| | | | | | 102 | 20 | SAMN40750186 |
| | | | | | 137 | 20 | SAMN40750187 |
| | | | | | 141 | 21.5 | SAMN40750188 |
| | | | | | 152 | NA | SAMN40750189 |
| 18323 | RDV | 74-78 | 131 | 21L | 0 | 26 | SAMN40750190 |
| | Paxlovid | 81-85 | | | 6 | 17.1 | SAMN40750191 |
| | | | | | 29 | 16.3 | SAMN40750192 |
| | | | | | 73 | NA | SAMN40750193 |
| 17072 | Paxlovid | 0-4 | 81 | 21K | 22 | NA | SAMN40750164 |
| | RDV | 20-24,<br>31-35,<br>63-67 | | | 52 | 21.9 | SAMN40750165 |
| | | | | | 68 | 20.2 | SAMN40750166 |
| | | | | | 77 | NA | SAMN40750167 |
| | | | | | 80 | 25.7 | SAMN40750168 |
| | | | | | 81 | NA | SAMN40750169 |

Note: [+]indicates that the patient died in the following month after hospitalization without retesting for SARS-CoV-2. [@]indicates that the patient died while SARS-CoV-2 positive. RDV-remdesivir. pd-post COVID-19 diagnosis. Ct-cycle threshold. NA-not available. Sequencing data are available under the BioProject PRJNA1088540.

**Supplementary Table 2.** Non-synonymous SARS-CoV-2 substitutions identified in nsp5 and nsp12. Samples are compared to their SARS-CoV-2 clade reference. Mutations exceeding 98% frequency of the sequence reads in all samples sequenced per patient may signify strain-specific diversity that established the infection, rather than fixation over the course of the infection.

| Patient ID | Total Collections Sequenced | Clade | Variant | Frequency Range | Number of Collections with Variant |
|---|---|---|---|---|---|
| 1995 | 3 | 20C | nsp12: A16V | 1 | 3 |
| 10939 | 2 | 20C | nsp5: P96S | 1 | 2 |
| 11595 | 4 | 20G | nsp12: E136A | 0.27 | 1 |
| | | | nsp12: V166L | 0.11 | 1 |
| | | | nsp12: Q444K | 0.05 - 0.07 | 2 |
| | | | nsp12: V792I | 0.09 - 0.31 | 3 |
| | | | nsp12: M794I | 0.17 - 0.9 | 3 |
| | | | nsp12: C799F | 0.1 | 1 |
| | | | nsp12: V820G | 0.03 | 1 |
| 12105 | 2 | 20G | nsp12: V605I | 1 | 2 |
| 16902 | 10 | 21K | nsp12: C464Y | 1 | 2 |
| | | | nsp12: C799Y | 0.98 | 1 |
| 16915 | 4 | 21K | nsp12: T644M | 0.06 - 0.07 | 2 |
| 17072 | 6 | 21K | nsp5: M165I | 0.09 | 1 |
| | | | nsp5: T169I | 0.34 - 0.92 | 4 |
| | | | nsp5: A173T | 0.1 - 0.74 | 2 |
| | | | nsp5: G283C | 0.05 | 1 |
| | | | nsp12: T643I | 0.26 | 1 |
| | | | nsp12: V792I | 0.27 - 0.98 | 4 |
| | | | nsp12: E796K | 0.23 - 1 | 3 |
| 17320 | 3 | 21K | nsp12: I171M | 0.05 | 1 |
| 17423 | 14 | 21K | nsp5: L253I | 0.03 - 0.04 | 2 |
| | | | nsp12: V820G | 0.03 | 1 |
| | | | nsp12: F859L | 0.03 | 1 |
| 18323 | 4 | 21L | nsp5: T21I | 0.04 | 1 |

**Supplementary Table 3.** Relevant laboratory data of patient 11595 during the disease course.

| **Laboratory Parameter** | **Days after COVID-19 Diagnosis** | | | | | | | | |
|---|---|---|---|---|---|---|---|---|---|
| | **Normal Range** | **0** | **7** | **14** | **20** | **23** | **26** | **30** | **38** |
| CT value | | | | 19.4 | 19.25 | | 20.7 | | 20.7 |
| WBC count, ($\times10^3$/uL) | 3.4-11.2 | 3.79 | 2.33 | 2.8 | 1.47 | 1.45 | 1.62 | 3.98 | 4.02 |
| Neutrophil count, ($\times10^3$/uL) | 1.8-7.0 | ** | ** | 2.38 | 1.22 | 0.85 | 1.38 | 3.0 | 3.59 |
| Neutrophils (%) | 45-75 | ** | ** | 85 | 83.1 | 82 | 81 | 93 | 89.4 |
| Lymphocyte count, ($\times10^3$/uL) | 1.18-3.74 | ** | ** | 0.22 | 0.13 | 0.08 | 0.16 | 0.08 | 0.08 |
| Lymphocytes (%) | 20-50 | ** | ** | 8 | 8.8 | 8 | 10 | 2 | 2 |
| D- Dimer (ng/mL) | 0-229 | ** | ** | ** | ** | 451 | ** | ** | ** |
| CRP (mg/dL) | <=0.9 | ** | ** | ** | ** | 19.0 | ** | ** | ** |
| ESR (mm/hr) | 0-20 | ** | 5 | ** | ** | 25 | ** | ** | ** |
| LDH (U/L) | 118-230 | ** | 321 | ** | 280 | 375 | 317 | 444 | ** |

Abbreviations. CRP: C-reactive protein; CT: cycle threshold; ESR: Erythrocyte sedimentation rate; LDH: Lactate dehydrogenase; WBC: White blood cells; ** Indicates that these laboratory values were not available.

**Supplementary Table 4.** Immune and serum profiles of patient 16902 during the disease course.

| Laboratory Parameter | Normal Range | Days after COVID-19 Diagnosis | | | | | | | | |
|---|---|---|---|---|---|---|---|---|---|---|
| | | 0 | 1 | 7 | 11 | 14 | 28 | 36 | 47 | 52 |
| CT value | | | | | | | 25.1 | | 23 | |
| WBC count, (×$10^3$/uL) | 3.4-11.2 | 1.0 | 1.22 | 5.8 | 2.2 | 2.8 | 3.53 | 4.17 | 1.03 | 1.63 |
| Neutrophil count, (×$10^3$/uL) | 1.56-6.13 | 0.6 | 0.83 | 4.2 | 1.5 | 2.1 | 2.93 | 3.59 | 0.83 | 1.55 |
| Neutrophils (%) | 45-75 | 61.4 | 66 | 80 | 66 | 80 | 83 | 86 | 79 | 95 |
| Lymphocyte count, (×$10^3$/uL) | 1.18-3.74 | 0.2 | 0.17 | 0.52 | 0.37 | 0.28 | 0.19 | 0.17 | 0.08 | 0.08 |
| Lymphocytes (%) | 20-50 | 18.5 | 14 | 9 | 16 | 10 | 5.4 | 4 | 8 | 6 |
| D- Dimer (ng/mL) | 0-229 | ** | ** | ** | ** | ** | 160 | 338 | 175 | 1588 |
| CRP (mg/dL) | <=0.9 | ** | 9.2 | ** | ** | ** | 7.9 | 0.7 | 0.4 | 0.4 |
| LDH (U/L) | 118-230 | 273 | 274 | 523 | 312 | 325 | 414 | 363 | 502 | 575 |
| Procalcitonin (ng/mL) | <=0.08 | ** | ** | ** | ** | ** | ** | ** | ** | ** |

Abbreviations. CRP: C-reactive protein; CT: cycle threshold; ESR: Erythrocyte sedimentation rate; LDH: Lactate dehydrogenase; WBC: White blood cells; ** Indicates that these laboratory values were not available.

**Supplementary Table 5.** Immune and serum profiles of patient 17072 during the disease course.

| Laboratory Parameter | Normal Range | -2 | 22 | 23 | 24 | 30 | 31 | 34 | 36 | 42 | 46 | 52 | 60 | 66 | 68 | 77 | 80 | 81 |
|---|---|---|---|---|---|---|---|---|---|---|---|---|---|---|---|---|---|---|
| CT value | | | | | | | | 24.4 | | | | 21.9 | 26.5 | | 20.2 | | 25.7 | |
| WBC count, ($\times 10^9$/L) | 3.4-11.2 | 10.5 | 14.12 | 18.36 | 15.4 | 21.22 | 12.86 | 21.79 | 23.53 | 15.57 | 15.23 | 15.72 | 14.18 | 9.6 | 8.82 | 14.43 | 11.48 | 14.04 |
| Neutrophil count, ($\times 10^9$/L) | 1.8-7.0 | 7.6 | 12.86 | 15.97 | 11.99 | 19.1 | 12.86 | 19.02 | 22.12 | 14.57 | 13.07 | 14.16 | 12.39 | 7.5 | 6.33 | 12.43 | 10.23 | 12.12 |
| Neutrophils (%) | 45-75 | 72.4 | 90 | 87 | 79.6 | 88 | 91.5 | 87.3 | 94.0 | 88.0 | 85.8 | 84.5 | 87.4 | 78.2 | 71.7 | 86.2 | 89.1 | 86.3 |
| Lymphocyte count, ($\times 10^9$/L) | 1.18-3.74 | 1.4 | 0.57 | 1.1 | 0.81 | 0.85 | 0.39 | 0.67 | 0.71 | 0.60 | 0.55 | 0.74 | 0.50 | 0.99 | 1.13 | 0.61 | 0.46 | 1.08 |
| Lymphocytes (%) | 20-50 | 13.1 | 3 | 6 | 5.4 | 4 | 2.7 | 3.1 | 3.0 | 3.6 | 3.6 | 4.5 | 3.5 | 10.3 | 12.8 | 4.2 | 4.0 | 7.7 |
| D- Dimer (ng/mL) | 0-229 | | <150 | | | <150 | | | | | | 161 | | 156 | | | | |
| CRP (mg/dL) | <=0.9 | | | 8.7 | 4.9 | 13.7 | | 14 | 20 | 11.3 | 9.1 | | | | | | | |
| ESR (mm/hr) | 0-20 | | 59 | | | 49 | | | | | | | | | | | | |
| LDH (U/L) | 118-230 | 183 | | | 289 | 386 | | | | | | | | | | | | |
| Procalcitonin (ng/mL) | <=0.08 | | 0.72 | | | 0.75 | | 0.49 | | 0.29 | | | | | | | 0.61 | |

Abbreviations. CRP: C-reactive protein; CT: cycle threshold; ESR: Erythrocyte sedimentation rate; IL-6: Interleukin-6; LDH: Lactate dehydrogenase; WBC: White blood cells; ** Indicates that no other laboratory values were available on the first day of SARS-CoV2 infection (Day 0).

**Supplementary Table 6.** Virus isolation from nasopharyngeal swabs of immunocompromised patients.

| Patient ID | Viral load (Ct value) | Days post-diagnosis | Virus isolation | Passages in Vero E6 TMPRSS2 | Consensus mutations (clinical sample vs prototype strain) | Consensus mutations in nsp5/nsp12 (clinical sample vs passage 3) | Clade |
|---|---|---|---|---|---|---|---|
| 11595 | 20.7 | 38 | yes | 3 | nsp12 M794I, Spike E484Q, Spike Q677H | nsp12 Q444K, nsp12 I794M, nsp12 M900V | 20G |
| 16902 | NA | 0 | yes | 3 | No mutations | No mutations | 21K |
| | NA | 1 | no | - | - | - | - |
| | NA | 7 | no | - | - | - | - |
| | NA | 11 | no | - | - | - | - |
| | NA | 18 | no | - | - | - | - |
| | 19.10 | 42 | no | - | - | - | - |
| | 25.4 | 50 | no | - | - | - | - |
| 17072 | 26.5 | 60 | no | - | - | - | - |
| | NA | 77 | yes | 3 | nsp5 T169I, nsp12 V223I, nsp12 V792I | No mutations | 21K |
| | 25.7 | 80 | no | - | - | - | - |
| | NA | 81 | yes | 3 | nsp5 T169I, nsp12 V223I, nsp12 V792I | nsp5 M165I, nsp5 I169T, nsp12 I792V | 21K |

Note: NA: Not available; - not available as no virus was isolated.

**Supplementary Table 7.** The $IC_{50}$ values (µM) of nirmatrelvir and remdesivir against SARS-CoV-2 isolates WT and $nsp5^{T169I}nsp12^{V792I}$ in this study.

| | Pre-treatment | | Simultaneous treatment | | Post-treatment | |
|---|---|---|---|---|---|---|
| Nirmatrelvir | $IC_{50}$ (µM) | Fold | $IC_{50}$ (µM) | Fold | $IC_{50}$ (µM) | Fold |
| WT | 1.77 | 1 | 0.95 | 1 | 1.67 | 1 |
| $nsp5^{T169I}nsp12^{V792I}$ | 3.93 | 2.22 | 2.89 | 3.04 | 3.62 | 2.17 |
| Remdesivir | $IC_{50}$ (µM) | Fold | $IC_{50}$ (µM) | Fold | $IC_{50}$ (µM) | Fold |
| WT | 8.8 | 1 | 6.68 | 1 | 7.5 | 1 |
| $nsp5^{T169I}nsp12^{V792I}$ | 16.7 | 1.9 | 15.29 | 2.34 | 14.56 | 1.94 |

**Supplementary Table 8.** Binding energies (in kcal/mol) of nirmatrelvir drug molecule against nsp5 protein of WT and nsp5$^{T169I}$nsp12$^{V792I}$ isolates. Hydrogen and hydrophobic interactions are highlighted, providing insights into the molecular interactions at the binding interface.

| | Binding Energy (kcal/mol) | H-bond | | | | Hydrophobic Interactions |
|---|---|---|---|---|---|---|
| | | Residue No. | No. of Bonds | Molecules | Bond Length ($A^0$) | |
| nsp5 of WT docked with nirmatrelvir | -7.8 | His163 | 2 | NE2-F3 | 3.16 | Met49, Leu141, Asn142, Ser144, Cys145, His164, Met165, Asp187, Arg188, Gln189, Gln192 |
| | | | | NE2-F2 | 3.17 | |
| | | Glu166 | 2 | N-O2 | 3.10 | |
| | | | | O-N2 | 3.25 | |
| nsp5 of nsp5$^{T169I}$nsp12$^{V792I}$ docked with nirmatrelvir | -6.9 | Gly143 | 1 | N-O1 | 3.01 | His41, Met49, Phe140, Leu141, Asn142, His164, Met165, Gln189 |
| | | Cys145 | 1 | SG-O1 | 3.03 | |
| | | Glu166 | 1 | N-O3 | 3.25 | |

**Supplementary Table 9**. Binding energies (in kcal/mol) of remdesivir drug molecules against nsp12 protein of WT and nsp5$^{T169I}$nsp12$^{V792I}$ isolates. Hydrogen and hydrophobic interactions are highlighted, providing insights into the molecular interactions at the binding interface.

<table>
<tr><th rowspan="2"></th><th rowspan="2">Binding Energy (kcal/mol)</th><th colspan="4">H-bond</th><th rowspan="2">Hydrophobic Interactions</th></tr>
<tr><th>Residue No.</th><th>No. of Bonds</th><th>Molecules</th><th>Bond Length ($A^0$)</th></tr>
<tr><td rowspan="10">nsp12 of WT docked with RTP</td><td rowspan="10">-7.1</td><td>Asp618</td><td>1</td><td>OD1-N5</td><td>3.13</td><td rowspan="10">Lys621, Ser681, Ser682, Thr687, Asp761,</td></tr>
<tr><td>Tyr619</td><td>1</td><td>O-O2</td><td>2.89</td></tr>
<tr><td>Cys622</td><td>1</td><td>N-O2</td><td>3.12</td></tr>
<tr><td rowspan="2">Asp623</td><td rowspan="2">2</td><td>OD1-O13</td><td>2.84</td></tr>
<tr><td>N-O13</td><td>3.03</td></tr>
<tr><td>Thr680</td><td>1</td><td>OG1-O9</td><td>2.86</td></tr>
<tr><td>Asn691</td><td>1</td><td>ND2-O6</td><td>2.81</td></tr>
<tr><td>Ser759</td><td>1</td><td>OG-O6</td><td>2.84</td></tr>
<tr><td rowspan="2">Asp760</td><td rowspan="2">2</td><td>OD2-O12</td><td>3.08</td></tr>
<tr><td>OD1-O12</td><td>3.29</td></tr>
<tr><td rowspan="5">nsp12 of nsp5$^{T169I}$nsp12$^{V792I}$ docked with RTP</td><td rowspan="5">-7.1</td><td rowspan="2">Asn497</td><td rowspan="2">2</td><td>N-O9</td><td>3.05</td><td rowspan="5">Val495, Asn496, Arg569, Gln573, Leu576, Thr686, Thr687, Gly683, Tyr689</td></tr>
<tr><td>O-O9</td><td>3.18</td></tr>
<tr><td>Ser682</td><td>1</td><td>O-O13</td><td>2.80</td></tr>
<tr><td>Asp684</td><td>1</td><td>O-O13</td><td>2.70</td></tr>
<tr><td>Ala685</td><td>1</td><td>O-O2</td><td>3.22</td></tr>
</table>

**Supplementary Table 10.** Primers used for whole-genome sequencing of SARS-CoV-2 on the Oxford Nanopore technologies GridION platform.

| Name | Sequence | Name | Sequence |
|---|---|---|---|
| 24 pool 1 | TTCTCCTAAGAAGCTATTAAAATCACATGG | 23 R pool 2 | TTCGCTGATTTTGGGGTCCA |
| 24 R pool 1 | GGCTCTTCCATATAGGCAGCT | 23 F pool 2 | TTTCCTCTGGCTGTTATGGC |
| 24 F pool 1 | TGGGTAGTCTTGTAGTGCGT | 21 R pool 2 alt_2 | TTGCAGCAGGATCCACAAGA |
| 22 R pool 1 | GCTGAGCCACATCAAGCCTA | 21 R pool 2 | AGCCAGCTATAAAACCTAGCCA |
| 22 F pool 1 | CCTCAATGAGGTTGCCAAGA | 21 F pool 2 alt_2 | TTTCAAACACGTGCAGGCTG |
| 20 R pool 1 | CTGCACCAAGTGACATAGTGT | 19 F pool 2 | ACAGATGCGCAAACAGGTTC |
| 20 F pool 1 | GGACCTTGAAGGAAAACAGGGT | 17 R pool 2 | TGTCACTACAAGGCTGTGCA |
| 18 R pool 1 | TGGCCATCTTTACACCAAAGC | 17 F pool 2 alt omi | TGTGTACATTGGCGACCCTG |
| 18 F pool 1 | TGCGGCTTGTAGAAAGGTTCA | 15 R pool 2 | GCCTCATAAAACTCAGGTTCCC |
| 16 R pool 1 | ACAATTTCAGCAGGACAACGC | 15 F pool 2 | ATGCACGCTGCTTCTGGTAA |
| 16 R pool 1 alt | AGGACAACGCCGACAAGTTC | 13 R pool 2 | GCAGACGGTACAGACTGTGT |
| 16 F pool 1 | TGCTTACCCACTTACTAAACATCCT | 13 R pool 2 alt | CCCACAGGGTCATTAGCACA |
| 16 F pool 1 alt | TGAACGGTTCGTGTCTTTAGC | 13 F pool 2 alt_2 | TCTTGTGCTGCCGGTACTAC |
| 14 R pool 1 | GCAGCATTACCATCCTGAGC | 11 R pool 2 alt_2 | TGGCTGCTGTTGTAAGAGGT |
| 14 F pool 1 | ATCCTTTGGTGGTGCATCGT | 11,855 F pool 2 | GTTGGGTGTTGGTGGCAAAC |
| 12 R pool 1_alt_2 | GCAAGTACAAACCTACCTCCCT | 11 F pool 2 alt | ATTGTTGGGTGTTGGTGGCA |
| 12 F pool 1 alt_2 | AATTTGACCGTGATGCAGCC | 9 R pool 2 alt omi | YCTCATAGCACATTGGTAAACAC |
| 10 R pool 1 | CTGGACACATTGAGCCCACA | 9 F pool 2 | TTTTGTCGTGCCTGGTTTGC |
| 10 F pool 1 | GACACCTAAGTATAAGTTTGTTCGC | 9 F pool 2 alt | GCCCATTGATTGCTGCAGTC |
| 8 R pool 1 | GCTGATGTTGCAAAGTCAGTGT | 7 F pool 2 alt omi | ACCAACCATATCCAAACGCA |
| 8 F pool 1 | GCCCCGATTTCAGCTATGGT | 7 R pool 2 alt omi | TGCAAAAGCCTTTACCTCCA |
| 6 R pool 1 | TCAATAGCCACCACATCACCA | 5 R pool 2 alt_2 | GTTCATACTGAGCAGGTGGTG |
| 6 R pool 1 alt | CAATAGCCACCACATCACCA | 5 F pool 2 alt_2 | ACGTGTTGAGGCTTTTGAGT |
| 6 F pool 1 alt_2 | GCTGTTATGTACATGGGCACAC | 3 R pool 2 | ACCGAGCAGCTTCTTCCAAA |
| 4 R pool 1 alt_2 | TGCTGACATGTACCTACCCAG | 3 F pool 2 | GTGAAGAAGAAGAGTTTGAGCCA |
| 4 F pool 1 | GGTGTGGTTGATTATGGTGCT | 1 R pool 2 | GACCTTCGGAACCTTCTCCA |
| 4 F pool 1 alt | GGGTGTGGTTGATTATGGTGCT | 1 F pool 2_alt_omi | ACCAACCAACTTTYGATCTCT |
| 2 R pool 1 | GCAGAAGTGGCACCAAATTCC | 19 R pool 2 omi | TCTACCAATGGTTCTAAAGCCG |
| 2 F pool 1 | TTCTTCGTAAGGGTGGTCGC | | |
